# The COVID-19 Trajectory in Israel: A Comprehensive Analysis of National Surveillance Data on Documented Infections, Test Positivity, and Vaccine-Effectiveness Estimates

**DOI:** 10.64898/2026.09.06.26362366

**Authors:** Yaakov Ophir

## Abstract

**Background:** Israel was among the first countries to implement nationwide BNT162b2 vaccination and became an important source of real-world evidence during the COVID-19 pandemic. The present study aimed to characterize the long-term national epidemic trajectory and the evolution of vaccination-associated differences in documented infection across the course of the pandemic.

**Methods:** Five Israeli Ministry of Health datasets were analyzed from February 2020 through May 2023. National trends in documented infection and test positivity were characterized. Vaccinated– unvaccinated differences were examined using routine-surveillance case rates. To reduce potential bias from differences in prior-infection history and real-world testing intensity, a second age-adjusted analysis compared test positivity among individuals without documented prior infection.

**Results:** The two completed pre-rollout waves declined to ≤10% of their peaks within 32 and 43 days, whereas the winter 2020–2021 wave, already underway when vaccination began, required 68 days. Delta and Omicron peaks were 17% and approximately ninefold higher than the January 2021 peak. Routine-surveillance apparent vaccine effectiveness peaked at 94.4% in February 2021, declined to 61.5% by late May, and remained negative from February 2022 onward. The age-adjusted test-positivity analysis among individuals without documented prior infection yielded generally lower estimates, which remained negative from December 2021 onward.

**Conclusions:** Overall, the early vaccination-associated advantage against documented infection was not stable over extended follow-up. While these observational findings cannot establish causal vaccine effects or address severe outcomes, they underscore the importance of evaluating vaccine-effectiveness estimates over extended follow-up as population immunity, testing practices, and viral variants evolve.

## Introduction

During the COVID-19 pandemic, Israel played a prominent role in the early evaluation of COVID-19 vaccine effectiveness under real-world conditions. The national rollout of the Pfizer–BioNTech BNT162b2 vaccine began on December 20, 2020, and its rapid implementation led Israel to be described as the “world’s laboratory” for the vaccination campaign (Birnhack, 2021). Within little more than two months, a large matched-cohort study using data from Clalit Health Services, Israel’s largest health fund, estimated 92% effectiveness against documented infection from 7 days after the second dose (Dagan et al., 2021). This was followed in May 2021 by a nationwide analysis of Ministry of Health surveillance data, which estimated approximately 95% effectiveness against SARS-CoV-2 infection during January–April 2021 (Haas et al., 2021).

The epidemiological picture, however, changed substantially over time. A renewed outbreak during the Delta period was accompanied by evidence that protection against infection waned with time since the second dose (Goldberg et al., 2021). A third dose was subsequently associated with a marked short-term reduction in confirmed infections relative to two-dose recipients (Bar-On et al., 2021). A longitudinal analysis of Israeli national data similarly documented changing vaccine-effectiveness patterns through the Delta period and booster rollout (Saban et al., 2021). During the Omicron period, protection against confirmed infection after a fourth dose was comparatively short-lived (Bar-On et al., 2022).

These changing estimates also highlight the difficulty of interpreting vaccine effectiveness from observational surveillance data, where vaccination is not randomly assigned and observed infection rates may be shaped by multiple time-varying factors, including epidemic dynamics, population behavior, public-health measures, and testing practices (Agampodi et al., 2024; Fung et al., 2024).

An important consideration is prior infection. Israeli data showed that individuals with documented previous SARS-CoV-2 infection had substantial protection against subsequent infection (Goldberg et al., 2022). As the epidemic progressed and prior infections accumulated, vaccination-status groups could therefore differ not only in vaccination history but also in the prevalence of infection-derived protection. Comparisons based on vaccination status alone may thus increasingly conflate vaccination status with differences in prior-infection history.

Another important consideration is differential testing by vaccination status. The nationwide Ministry of Health analysis described above addressed this issue directly, stating: “*Israel’s SARS-CoV-2 testing policy was different for unvaccinated and vaccinated individuals…At 7 days after the second dose, vaccinated individuals were exempt from the SARS-CoV-2 testing required of individuals who either had contact with a laboratory-confirmed case or returned from travel abroad. This testing policy might have resulted in a differential bias that would cause overestimation of vaccine effectiveness against asymptomatic infection (ie, asymptomatic people who received two doses were less likely to be tested than unvaccinated asymptomatic people)*” (Haas et al., 2021).

These issues are especially relevant when vaccine-effectiveness estimates are considered over the course of the epidemic rather than within a single period. Most Israeli vaccine-effectiveness studies examined specific phases of the epidemic, vaccination schedules, or variant periods. Yet the national surveillance record spans the full sequence of pre-vaccination waves, the primary vaccination campaign, booster rollout, Delta, and Omicron. Over this period, vaccination status, accumulated prior infection, circulating variants, and testing practices changed concurrently. Examining the complete national trajectory therefore provides an opportunity to compare the pattern apparent in routine vaccination-status surveillance with the pattern observed when documented prior infection and testing are incorporated explicitly.

The present study examines the national COVID-19 trajectory in Israel from February 2020 through May 2023 using five Ministry of Health surveillance datasets. We first characterize the overall trajectory of documented infections and test positivity before and after vaccine rollout. We then compare the vaccine-effectiveness pattern apparent in routine surveillance with an age-adjusted analysis that incorporates documented prior infection and testing. Importantly, the present study does not aim to derive a causal estimate of vaccine effectiveness, but rather to determine how the observed vaccination-associated contrast evolved across the epidemic and how sensitive it is to prior-infection composition and testing ascertainment. Understanding how these factors shape long-term surveillance estimates may help improve the interpretation and design of vaccine-effectiveness monitoring in future epidemics and vaccination campaigns.

## Methods

### Study design and data sources

We conducted a retrospective longitudinal analysis of publicly available, aggregate national COVID-19 surveillance data from the Israeli Ministry of Health (IMOH), spanning the beginning of the epidemic through May 27, 2023. This endpoint fell 22 days after the World Health Organization determined that COVID-19 no longer constituted a Public Health Emergency of International Concern (WHO, 2023).

Five datasets are provided as Supplementary Datasets S1–S5. Datasets S1–S3 were obtained from the IMOH *World of Data* COVID-19 dashboard (IMOH, 2026). Dataset S1 (*Daily Confirmed Cases*) contains daily confirmed SARS-CoV-2 infections from February 12, 2020; Dataset S2 (*Daily Test Positivity*) contains the daily percentage of tested individuals who were positive from February 20, 2020; and Dataset S3 (*Daily Cases and Rates by Vaccination Status*) contains daily case counts and rates per 100,000 population from January 17, 2021, reported for all ages and separately for persons aged <60 and ≥60 years and stratified as vaccinated, vaccination status expired, or unvaccinated. All analyses of S1–S3 were truncated at May 27, 2023.

Datasets S4 and S5 were obtained from the IMOH *COVID-19 Database* on the Israel Government Data portal (IMOH, 2023). Dataset S4 (*Weekly Cases by Vaccination, Age, and Recovery Status*) reports weekly confirmed cases by age, vaccination status, and previous documented infection; Dataset S5 (*Weekly Tests by Vaccination, Age, and Recovery Status*) reports corresponding numbers tested by age, previous documented infection, and vaccine dose category. Unlike S1–S3, these weekly datasets begin later, on May 30, 2021, but extend through the same study endpoint of May 27, 2023. Accordingly, the testing- and prior-infection–informed analysis could begin only on May 30, 2021, the first week for which case and testing data with the required vaccination and prior-infection stratification were jointly available.

For dissemination, Hebrew headings and recurring labels in S1–S3 were translated into English without altering numerical values, dates, missing values, or privacy-suppressed cells. S4 and S5 were supplied with English variable names. Processing conventions are documented in the accompanying README.

### National epidemic trajectory

The overall epidemic trajectory was characterized independently of vaccination status using S1 and S2. Confirmed infections were examined using the source-provided 7-day moving average, and a 7-day moving average was calculated for overall test positivity.

To compare wave resolution descriptively, we examined the two completed major waves preceding vaccine rollout and the winter 2020–2021 wave spanning its initiation. For each wave, the peak was defined as the maximum 7-day moving-average case count, and resolution was measured as the number of days from the peak to the first subsequent day at which the moving average reached ≤50%, ≤25%, and ≤10% of the peak. The smaller July 2020 local maximum was treated as part of the broader summer–autumn 2020 wave because it was followed by only a partial decline before the larger September peak. Subsequent major peak magnitudes were also compared descriptively. The calendar windows used to identify the named epidemic waves are specified in the reproducibility code.

### Vaccination-status analyses

Dataset S3 was used to reconstruct the vaccinated–unvaccinated pattern visible in routine IMOH surveillance. The primary analysis used the *All ages* category; secondary analyses examined persons aged <60 and ≥60 years.

Individuals classified as vaccinated and vaccination-status-expired were combined. Because the published rates for these categories use different population denominators, category-specific daily denominators were reconstructed as:

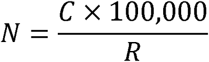

where *C* is the reported number of cases and *R* is the reported case rate per 100,000. Denominators were reconstructed only when both case counts and reported rates were non-zero. Because rates were rounded to one decimal place, weekly denominators were represented by the median of available daily estimates, while weekly cases were summed. Calendar weeks were defined from Sunday through Saturday.

The vaccinated and vaccination-status-expired denominators and cases were then combined, and weekly documented-infection rates were recalculated for the combined vaccinated and unvaccinated groups. The resulting risk ratio was expressed on the conventional vaccine-effectiveness scale:

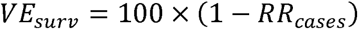

Negative values were retained. We refer to this measure as *apparent surveillance vaccine effectiveness*. Dataset S3 uses the IMOH routine vaccination-status definitions, which are not identical to the dose-count categories in S4 and S5; the two analyses were therefore compared as longitudinal surveillance trajectories rather than as estimates from identical cohorts.

### Testing- and prior-infection–informed analysis

Datasets S4 and S5 were harmonized by calendar week and age. S4 uses eight age groups (0– 19, 20–29, 30–39, 40–49, 50–59, 60–69, 70–79, and ≥80 years). In S5, ages 0–9 and 10–19 were combined to form 0–19, and ages 80–89 and ≥90 were combined to form ≥80.

The primary comparison was restricted to individuals without a previous documented infection. In S4, vaccinated cases were defined as Not_recovered_at_least_2_doses and unvaccinated cases as Not_recovered_not_vaccinated. Because the vaccinated case category includes recipients of two or more doses, the corresponding S5 testing denominator combined Non_recovered_2_doses and Non_recovered_3_and_more_doses; the unvaccinated denominator was Non_recovered_not_vaccinated. One-dose recipients were excluded.

Within each week and age stratum, test positivity was calculated as, where is the number of confirmed cases in S4 and the corresponding number tested in S5. Vaccinated-to-unvaccinated positivity risk ratios were expressed on the same VE scale:

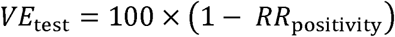

For the primary all-age trajectory, weekly Mantel–Haenszel risk ratios were calculated across the eight harmonized age strata. Secondary estimates for persons aged <60 and ≥60 years used the finer age strata within each broad group. Ninety-five percent confidence intervals were calculated on the log scale using the Greenland–Robins variance estimator.

Before linking S4 and S5, internal consistency was assessed by aggregating S5 to the S4 age categories and comparing total weekly numbers tested with the Tests_num field in S4. The harmonized totals were closely aligned: across 832 matched week-by-age cells, the median S5-to-S4 ratio was 0.988, the median absolute relative difference was 1.19%, and 96.9% of cells differed by no more than 5%.

The primary longitudinal comparison contrasted the S3 *All ages* apparent-surveillance trajectory with the age-adjusted S4–S5 trajectory in terms of direction, magnitude, and timing. Across overlapping weeks, we also summarized the number of weeks in which each estimate was positive or negative and, among weeks with a positive routine-surveillance estimate, compared the paired estimates and their median values. Calendar time was the underlying time scale; variant periods, vaccination milestones, and major testing-policy changes were included only as contextual figure annotations and were not modeled as causal exposures.

### Sensitivity analyses

Privacy-suppressed cells reported as <5 were assigned 2.5 in the primary analysis and values of 1 and 4 in sensitivity analyses. Additional analyses (1) restricted the S4–S5 comparison to adults aged ≥20 years and (2) repeated S3 denominator reconstruction using only observations with at least 5 or at least 10 cases in the corresponding vaccination-status category. For the full-period sensitivity summaries, Mantel–Haenszel strata were defined jointly by age group and calendar week.

## Results

### National epidemic trajectory

The national surveillance series showed successive waves of documented SARS-CoV-2 infection between February 2020 and May 2023 (Figure 1). Before the vaccination campaign began on December 20, 2020, two major waves had been completed. The first peaked on April 6, 2020, at a 7-day moving average of 616 confirmed cases per day, and the second on September 28 at 6,283 cases per day. A smaller local maximum in July 2020 occurred within the broader summer–autumn wave.

**Figure 1.**
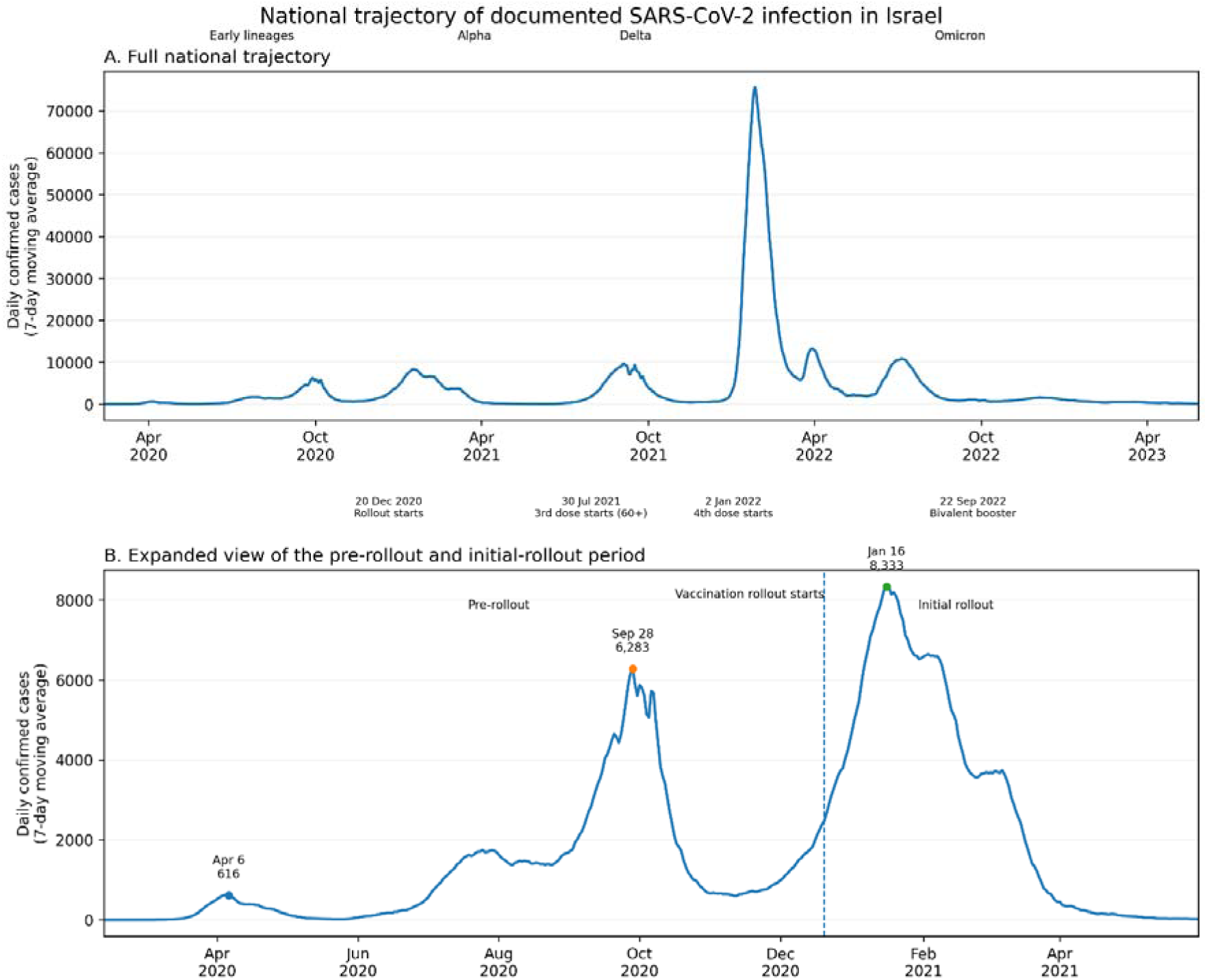
Note: Panel A shows the full national trajectory from February 12, 2020 through May 27, 2023, using the 7-day moving average of daily confirmed cases. Panel B provides an expanded view of February 12, 2020 through May 31, 2021, highlighting the two completed pre-rollout waves and the winter 2020–2021 wave spanning the start of the vaccination campaign. Contextual annotations indicate approximate periods of variant predominance and major vaccination milestones; these annotations are descriptive and were not modeled as independent causal exposures.

Following the April and September peaks, the 7-day moving average fell to ≤50% of the peak within 15 days in both waves, to ≤25% within 25 and 22 days, respectively, and to ≤10% within 32 and 43 days. By the start of vaccine rollout, the subsequent winter wave was already rising, with a 7-day moving average of 2,475 cases per day, approximately 30% of its eventual peak. It peaked 27 days later, on January 16, 2021, at 8,333 cases per day. The corresponding times to ≤50%, ≤25%, and ≤10% of the peak were 33, 60, and 68 days.

Subsequent major peaks were not uniformly smaller. The Delta-period wave peaked at 9,746 cases per day on September 4, 2021, approximately 17% above the January 2021 peak. The largest wave occurred during Omicron, reaching 75,624 cases per day on January 26, 2022, approximately 9.1 times the January 2021 peak. Further peaks of 13,321 and 10,861 cases per day occurred on March 30 and July 5, 2022, respectively.

Overall test positivity also varied markedly over the study period, with repeated increases corresponding to major epidemic waves and its highest levels occurring during 2022 (Figure 2).

**Figure 2.**
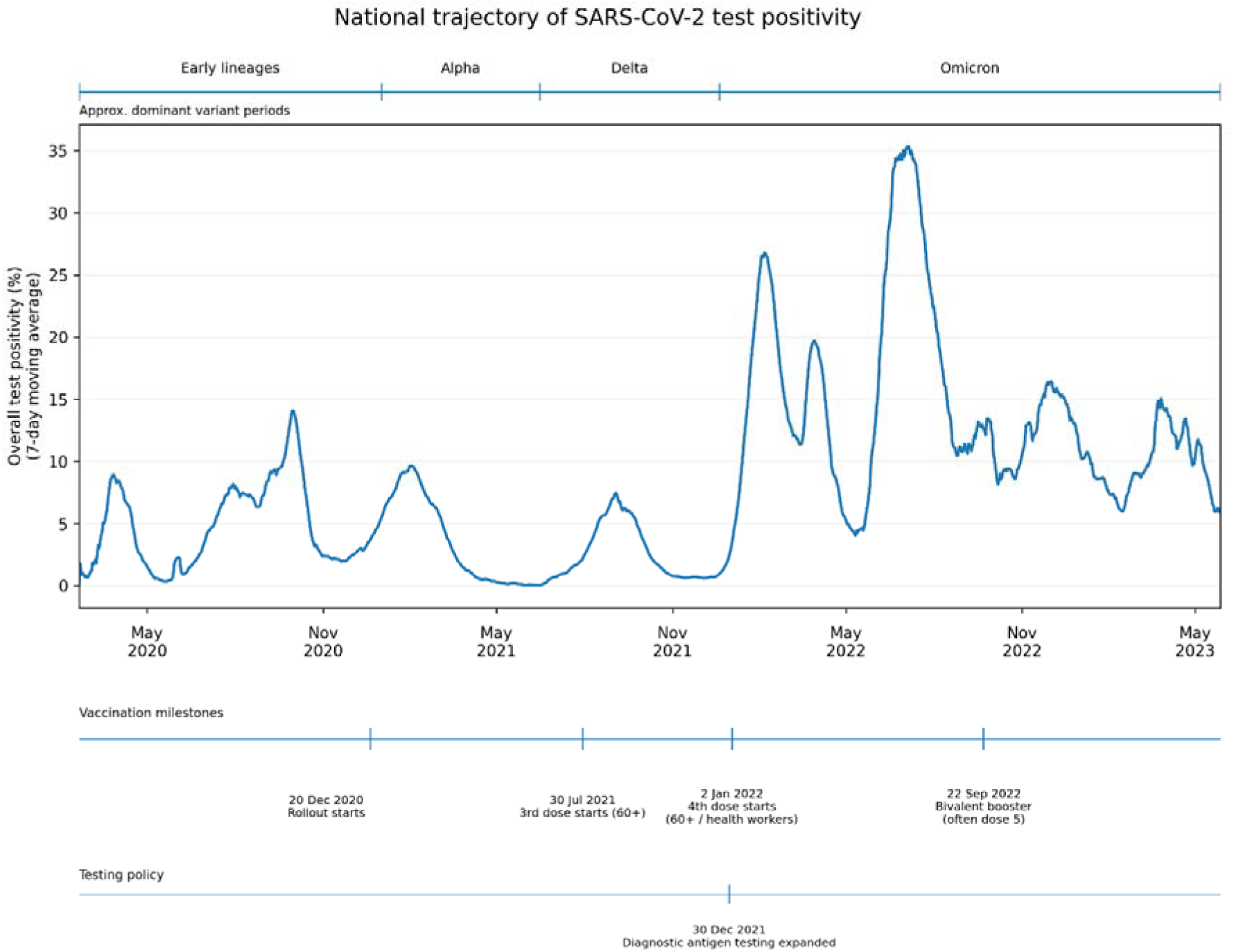
Note: The curve shows the 7-day moving average of overall test positivity from February 20, 2020 through May 27, 2023. Contextual annotations indicate approximate periods of variant predominance, major vaccination milestones, and the expansion of diagnostic antigen testing.

### Vaccination-associated trajectories

Routine surveillance initially showed a large difference in documented-infection rates between vaccinated and unvaccinated populations (Figure 3). Apparent surveillance vaccine effectiveness reached 94.4% in the week beginning February 14, 2021. At the beginning of the period in which the routine-surveillance and testing- and prior-infection–informed trajectories could be compared (Datasets S3 and S4–S5, respectively), the apparent surveillance estimate was 61.5% in the week beginning May 30, 2021. It subsequently increased to 72.3% on September 12 and 79.9% on December 5, before declining rapidly to 37.4% on December 19 and 8.2% one week later. The estimate first crossed below zero in the week beginning January 2, 2022 (−3.7%), returned temporarily to modestly positive values, and became negative again in the week beginning February 13, remaining below zero through the end of follow-up.

**Figure 3.**
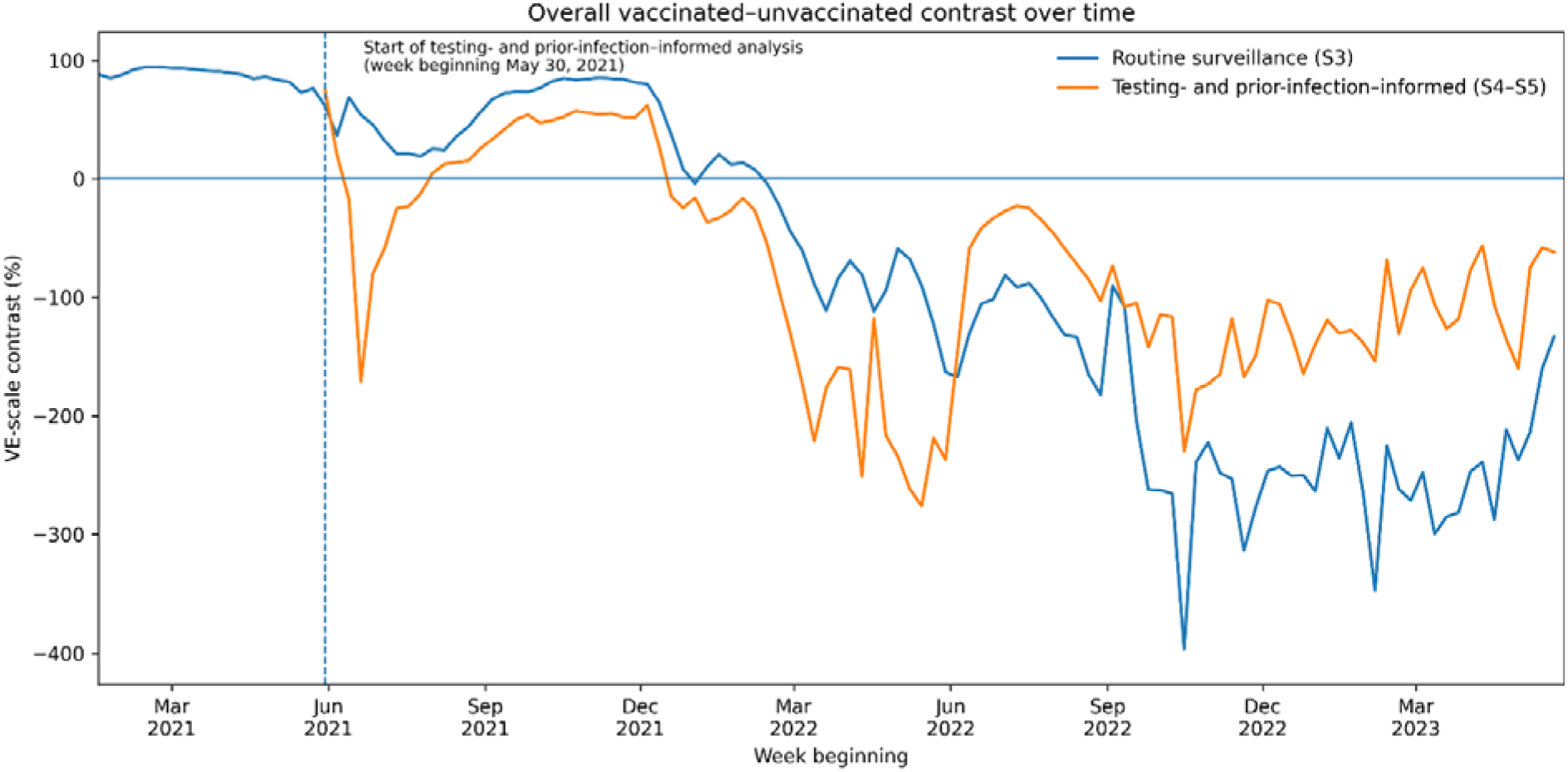
Note: The routine-surveillance trajectory is based on Dataset S3 and compares documented-infection rates in the combined vaccinated group with those in the unvaccinated group from January 17, 2021. The testing- and prior-infection–informed trajectory is based on Datasets S4–S5 and compares age-adjusted test positivity among vaccinated and unvaccinated individuals without previous documented infection from May 30, 2021, the first week for which the required case and testing data were jointly available. Both measures are shown on the VE scale; values below zero indicate higher observed infection rates or test positivity among vaccinated individuals.

The age-adjusted comparison restricted to individuals without a previous documented infection and to those who entered the testing system showed a different trajectory (Figure 3). In the first available week, beginning May 30, 2021, the Mantel–Haenszel RR was 0.250 (95% CI, 0.151– 0.414), corresponding to 75.0% on the VE scale (95% CI, 58.6%–84.9%). After unstable estimates during the low-incidence weeks of June and July, the contrast was positive from August through mid-December, including 41.6% in the week beginning September 12 and 62.1% on December 5. It crossed below zero earlier than the routine-surveillance estimate, reaching −14.5% in the week beginning December 19 (RR=1.145, 95% CI, 1.086–1.208), and remained negative in every subsequent week through May 27, 2023.

Across the 104 overlapping weeks, routine surveillance produced a positive all-age estimate in 36 weeks (Figure 3). The testing- and prior-infection–informed estimate was lower than the corresponding routine-surveillance estimate in 35 of these 36 weeks and was already negative in 14. Among the 36 weeks, the median estimates were 55.1% and 17.6%, respectively.

### Secondary and sensitivity analyses

Secondary age-stratified analyses showed the same broad transition. In the week beginning September 12, 2021, testing- and prior-infection–informed estimates were 38.9% among persons aged <60 and 73.4% among those aged ≥60. By December 19, the corresponding estimates had declined to −15.1% and −5.7%, respectively, and by March 27, 2022, to −149.6% and −232.4%.

The principal longitudinal findings were robust across sensitivity analyses. Assigning privacy-suppressed cells values of 1 or 4 rather than 2.5 produced a median absolute change of 0.0 percentage points in weekly estimates. Restricting the testing-informed analysis to adults aged ≥20 years preserved the late-2021 reversal and the predominantly negative pattern during 2022; the age- and week-stratified VE-scale estimate across the full period was −36.4% in the primary analysis and −41.9% after excluding persons aged <20 years. Restricting Dataset S3 denominator reconstruction to observations with at least 5 or at least 10 cases likewise produced a median absolute difference of 0.0 percentage points and did not alter the timing or direction of the principal transition.

## Discussion

The comprehensive analysis of Israel’s national surveillance record yielded three main findings. First, the national case series showed no sustained reduction in the magnitude of documented-infection waves during the post-rollout period. The winter 2020–2021 wave, which spanned the start of the vaccination campaign, also did not resolve more rapidly than the two completed pre-rollout waves. Second, the large early vaccinated–unvaccinated difference apparent in routine surveillance attenuated substantially over time and eventually reversed. Third, this attenuation occurred earlier and was generally more pronounced when the comparison was restricted to individuals without documented prior infection and to those who entered the testing system.

Importantly, these observational findings should not be interpreted as causal estimates of the effect of vaccination on the national epidemic trajectory. The counterfactual course of the epidemic without vaccination cannot be observed, and vaccination unfolded alongside multiple factors that could also influence infection patterns, including viral evolution, accumulated infection-derived immunity, public-health measures, behavioral changes, seasonality, and changes in testing.

Nevertheless, the findings place the high early Israeli vaccine-effectiveness estimates in a broader temporal context. Studies conducted during the initial campaign reported large reductions in documented infection among vaccinated populations (Dagan et al., 2021; Haas et al., 2021), while subsequent studies documented waning protection and renewed short-term protection following additional doses (Bar-On et al., 2021, 2022; Goldberg et al., 2021). These studies addressed important questions within particular epidemiological periods. The present analysis complements this literature by placing these period-specific estimates within the full national epidemic trajectory.

Viewed across the epidemic as a whole, the large early advantage against documented infection attenuated substantially and eventually reversed. The later negative VE-scale values should be interpreted on the same observational footing as the earlier positive values: they indicate that the observed vaccinated–unvaccinated contrast had reversed, but do not by themselves explain why.

Waning protection and immune escape can explain movement toward zero, but not a reversal below zero. Such a reversal could reflect a genuine difference in susceptibility, potentially involving altered immune responses following repeated vaccination. Alternatively, it could arise from remaining differences between the groups, including undocumented prior infection, differences in who was tested, and social exposure shaped by behavior or vaccination-related policies.

These alternative explanations underscore the importance of addressing, where possible, key sources of bias in routine surveillance data. The second analysis was designed to reduce potential bias from previous infection, which can reduce subsequent infection risk, and differences in testing, which can affect the probability that infections are detected. In this analysis, the vaccination-associated contrast attenuated earlier and was generally lower than in routine surveillance. The divergence between the two trajectories illustrates how surveillance-based vaccine-effectiveness estimates can depend on the composition of the groups being compared and on the testing process through which infections are observed.

### Limitations and Conclusions

Several limitations should guide interpretation. The analyses rely on aggregate surveillance data, limiting adjustment for individual-level confounding and causal attribution. Prior infection was captured only when documented; restriction to tested individuals does not eliminate selection into testing; vaccination categories differed somewhat across datasets; and calendar time encompassed concurrent changes in variant circulation, time since vaccination, booster uptake, behavior, and testing. The study also concerns documented infection and test positivity only, not hospitalization, severe disease, or death.

Within these limits, the national surveillance record indicates that the early vaccination-associated advantage against documented infection was not sustained across the evolving epidemic: the vaccinated–unvaccinated contrast attenuated and eventually reversed, while major post-rollout waves remained substantial and did not show a consistent pattern of more rapid resolution. The magnitude of vaccination-associated contrasts was also sensitive to prior-infection composition and testing. Future surveillance would therefore benefit from routinely linking vaccination status with prior-infection history, age, time since vaccination, and testing denominators using stable definitions over time. The Israeli experience thus provides a case study of how short-term effectiveness estimates and longer-term population-level infection patterns can diverge as an epidemic evolves.

## Supplementary Materials

Datasets S1–S5, an accompanying README describing the datasets and processing conventions, and Python code for reproducing the analyses and results.

## Author Contributions

Conceptualization: YO

Methodology: YO and GS

Data Curation: YO and GS

Formal Analysis: YO and GS

Validation: GS

Investigation: YO

Visualization: YO

Writing – Original Draft: YO

Writing – Review & Editing: YO and GS

Project Administration: YO

## Declarations

### Ethics

The study was approved by the Ariel University Ethics Committee (approval no. AU-SOC-YO-20260906). It used publicly available aggregate surveillance data and involved no individual-level records, participant contact, or individual-level data linkage.

### Data and Code Availability

The five source datasets, a README describing their contents and processing conventions, and the complete reproducibility code are provided as supplementary materials.

### AI usage

Generative AI tools were used to assist with language editing, manuscript refinement, and reproducibility code. The authors take full responsibility for the analyses and scientific content.

## Data Availability

All data analyzed in this study and the complete reproducibility code are provided as supplementary materials. These include the five source datasets and a README describing their contents and processing conventions.

