## Supplementary material for "The COVID-19 Trajectory in Israel: A Comprehensive Analysis of National Surveillance Data on Documented Infections, Test Positivity, and Vaccine-Effectiveness Estimates": Ophir_Reproduce_Code_Israel_Covid_Analysis.py

```
#!/usr/bin/env python3
"""
Reproducibility script for:

    The COVID-19 Trajectory in Israel: A Comprehensive Analysis of
    National Surveillance Data on Documented Infections, Test Positivity,
    and Vaccine-Effectiveness Estimates

This script starts from the five Supplementary Datasets (S1-S5) and
reproduces the analyses reported in the manuscript Results section.

The script is deliberately written as a transparent, step-by-step workflow.
Each numbered section corresponds to a component of the Methods/Results:

    1. Load and validate the five source datasets
    2. Characterize the national epidemic trajectory (S1-S2)
    3. Reconstruct routine-surveillance apparent VE (S3)
    4. Harmonize and link weekly case/testing data (S4-S5)
    5. Calculate testing- and prior-infection-informed estimates (S4-S5)
    6. Compare the two longitudinal trajectories
    7. Run the prespecified sensitivity analyses
    8. Verify the manuscript-facing numerical results
    9. Export analysis tables, summary text, and figures

No random procedures are used.

Expected input filenames
------------------------
Dataset_S1_Daily_Confirmed_Cases_Israel.xlsx
Dataset_S2_Daily_Test_Positivity_Israel.xlsx
Dataset_S3_Daily_Cases_and_Rates_by_Vaccination_Status_Israel.xlsx
Dataset_S4_Weekly_Cases_by_Vaccination_Age_and_Recovery_Status_Israel.csv
Dataset_S5_Weekly_Tests_by_Vaccination_Age_and_Recovery_Status_Israel.csv

Usage
-----
Place this script in the same folder as S1-S5 and run:

    python reproduce_israel_covid_analysis.py

or specify folders explicitly:

    python reproduce_israel_covid_analysis.py \
        --data-dir /path/to/data \
        --output-dir /path/to/output

Dependencies
------------
Python 3.10+
pandas
numpy
matplotlib
openpyxl
"""

from __future__ import annotations

import argparse
import hashlib
import math
from pathlib import Path
from statistics import NormalDist
from typing import Iterable

import matplotlib.pyplot as plt
import matplotlib.dates as mdates
import numpy as np
import pandas as pd


# =============================================================================
# 0. Analysis constants
# =============================================================================

STUDY_END = pd.Timestamp("2023-05-27")
ROLLOUT_START = pd.Timestamp("2020-12-20")
LINKED_START = pd.Timestamp("2021-05-30")
LINKED_LAST_WEEK = pd.Timestamp("2023-05-21")  # week ending 2023-05-27
SUPPRESSED_PRIMARY = 2.5
Z_975 = NormalDist().inv_cdf(0.975)

FILES = {
    "S1": "Dataset_S1_Daily_Confirmed_Cases_Israel.xlsx",
    "S2": "Dataset_S2_Daily_Test_Positivity_Israel.xlsx",
    "S3": "Dataset_S3_Daily_Cases_and_Rates_by_Vaccination_Status_Israel.xlsx",
    "S4": "Dataset_S4_Weekly_Cases_by_Vaccination_Age_and_Recovery_Status_Israel.csv",
    "S5": "Dataset_S5_Weekly_Tests_by_Vaccination_Age_and_Recovery_Status_Israel.csv",
}

AGE_ORDER = [
    "0-19", "20-29", "30-39", "40-49",
    "50-59", "60-69", "70-79", "80+",
]
AGES_LT60 = ["0-19", "20-29", "30-39", "40-49", "50-59"]
AGES_60PLUS = ["60-69", "70-79", "80+"]
AGES_ADULT = ["20-29", "30-39", "40-49", "50-59", "60-69", "70-79", "80+"]

EXPECTED_S5_AGE_LABELS = {
    "0-9", "10-19", "20-29", "30-39", "40-49",
    "50-59", "60-69", "70-79", "80-89", "90+",
}

# Windows used only to locate the descriptive peaks named in the manuscript.
# They are not used in any vaccine-effectiveness calculation.
WAVE_WINDOWS = {
    "First pre-rollout wave": ("2020-03-01", "2020-05-31"),
    "Summer-autumn 2020 wave": ("2020-06-01", "2020-11-30"),
    "Winter 2020-2021 wave": ("2020-12-01", "2021-05-31"),
    "Delta-period wave": ("2021-06-01", "2021-12-15"),
    "Omicron BA.1-period wave": ("2021-12-15", "2022-03-15"),
    "Spring 2022 wave": ("2022-03-16", "2022-05-31"),
    "Summer 2022 wave": ("2022-06-01", "2022-08-31"),
}

# Contextual dates used only for figure annotations. They are not analytic
# exposures and do not enter any estimate.
VARIANT_BOUNDARIES = [
    (pd.Timestamp("2021-01-01"), "Alpha"),
    (pd.Timestamp("2021-06-15"), "Delta"),
    (pd.Timestamp("2021-12-20"), "Omicron"),
]
VACCINATION_MILESTONES = [
    (pd.Timestamp("2020-12-20"), "Vaccination rollout starts"),
    (pd.Timestamp("2021-07-30"), "3rd dose starts (60+)"),
    (pd.Timestamp("2022-01-02"), "4th dose starts"),
    (pd.Timestamp("2022-09-22"), "Bivalent booster offered"),
]
TESTING_MILESTONES = [
    (pd.Timestamp("2021-12-30"), "Diagnostic antigen testing expanded"),
]


# =============================================================================
# 1. Generic helpers
# =============================================================================

def require_columns(df: pd.DataFrame, columns: Iterable[str], dataset: str) -> None:
    """Fail early if a source dataset is not the expected version."""
    missing = [c for c in columns if c not in df.columns]
    if missing:
        raise ValueError(f"{dataset} is missing required columns: {missing}")


def sha256(path: Path) -> str:
    """Return a SHA-256 checksum for provenance logging."""
    h = hashlib.sha256()
    with path.open("rb") as f:
        for block in iter(lambda: f.read(1024 * 1024), b""):
            h.update(block)
    return h.hexdigest()


def parse_week_start(series: pd.Series) -> pd.Series:
    """Parse 'YYYY-MM-DD - YYYY-MM-DD' and return the Sunday week start."""
    return pd.to_datetime(series.astype(str).str.split(" - ").str[0], errors="raise")


def sunday_week_start(date_series: pd.Series) -> pd.Series:
    """Map daily dates to Sunday-Saturday calendar weeks."""
    return date_series.dt.to_period("W-SAT").dt.start_time


def parse_suppressed_count(series: pd.Series, suppressed_value: float) -> pd.Series:
    """Convert counts to numeric, assigning '<5' the requested value."""
    cleaned = (
        series.astype(str)
        .str.strip()
        .replace({"<5": str(suppressed_value), "nan": np.nan, "": np.nan})
    )
    return pd.to_numeric(cleaned, errors="coerce")


def fmt_date(value: pd.Timestamp) -> str:
    return pd.Timestamp(value).strftime("%Y-%m-%d")


def assert_close(label: str, observed: float, expected: float, atol: float = 1e-8) -> None:
    if not math.isclose(float(observed), float(expected), abs_tol=atol, rel_tol=0.0):
        raise AssertionError(f"{label}: observed={observed}, expected={expected}")


def get_week_row(df: pd.DataFrame, date_string: str) -> pd.Series:
    week = pd.Timestamp(date_string)
    rows = df.loc[df["week_start"].eq(week)]
    if len(rows) != 1:
        raise AssertionError(f"Expected exactly one row for {date_string}; found {len(rows)}")
    return rows.iloc[0]


# =============================================================================
# 2. Load and validate Supplementary Datasets S1-S5
# =============================================================================

def load_datasets(data_dir: Path):
    paths = {key: data_dir / filename for key, filename in FILES.items()}
    missing = [str(path) for path in paths.values() if not path.exists()]
    if missing:
        raise FileNotFoundError(
            "The following required input files were not found:\n" + "\n".join(missing)
        )

    # S1-S3 contain a worksheet title in row 1; row 2 contains the true header.
    s1 = pd.read_excel(paths["S1"], header=1, engine="openpyxl")
    s2 = pd.read_excel(paths["S2"], header=1, engine="openpyxl")
    s3 = pd.read_excel(paths["S3"], header=1, engine="openpyxl")
    s4 = pd.read_csv(paths["S4"], dtype=str, encoding="utf-8-sig")
    s5 = pd.read_csv(paths["S5"], dtype=str, encoding="utf-8-sig")

    require_columns(
        s1,
        ["Date", "Daily_confirmed_cases", "7_day_moving_average", "Cumulative_confirmed_cases"],
        "Dataset S1",
    )
    require_columns(
        s2,
        ["Date", "PCR_positivity_percent", "Institutional_antigen_positivity_percent", "Overall_positivity_percent"],
        "Dataset S2",
    )
    require_columns(
        s3,
        [
            "Date", "Age_group",
            "Cases_vaccinated", "Cases_expired_vaccination_status", "Cases_unvaccinated",
            "Cases_per_100k_vaccinated", "Cases_per_100k_expired_vaccination_status",
            "Cases_per_100k_unvaccinated",
        ],
        "Dataset S3",
    )
    require_columns(
        s4,
        [
            "Week", "Age_group", "Tests_num",
            "Not_recovered_at_least_2_doses", "Not_recovered_partially_vaccinated",
            "Not_recovered_not_vaccinated", "Cases_among_recoverd_with_vacc",
            "Cases_among_recovered_without_vacc",
        ],
        "Dataset S4",
    )
    require_columns(
        s5,
        [
            "Test_week", "Age_group",
            "Recovered_not_vaccinated", "Recovered_1_dose", "Recovered_2_doses",
            "Recovered_3_and_more_doses", "Non_recovered_not_vaccinated",
            "Non_recovered_1_dose", "Non_recovered_2_doses",
            "Non_recovered_3_and_more_doses",
        ],
        "Dataset S5",
    )

    # The released S5 file should contain the literal age label '10-19'.
    # This strict validation prevents spreadsheet date auto-formatting from
    # silently altering the category.
    observed_s5_ages = set(s5["Age_group"].astype(str).str.strip().dropna().unique())
    if observed_s5_ages != EXPECTED_S5_AGE_LABELS:
        raise ValueError(
            "Unexpected Age_group labels in Dataset S5.\n"
            f"Observed: {sorted(observed_s5_ages)}\n"
            f"Expected: {sorted(EXPECTED_S5_AGE_LABELS)}"
        )

    # Parse dates/numeric values in S1-S3.
    for df in (s1, s2, s3):
        df["Date"] = pd.to_datetime(df["Date"], format="%d-%m-%Y", errors="raise")

    for col in ["Daily_confirmed_cases", "7_day_moving_average", "Cumulative_confirmed_cases"]:
        s1[col] = pd.to_numeric(s1[col], errors="coerce")
    for col in ["PCR_positivity_percent", "Institutional_antigen_positivity_percent", "Overall_positivity_percent"]:
        s2[col] = pd.to_numeric(s2[col], errors="coerce")
    for col in s3.columns:
        if col not in {"Date", "Age_group"}:
            s3[col] = pd.to_numeric(s3[col], errors="coerce")

    # Restrict all daily datasets to the common study endpoint.
    s1 = s1.loc[s1["Date"] <= STUDY_END].sort_values("Date").reset_index(drop=True)
    s2 = s2.loc[s2["Date"] <= STUDY_END].sort_values("Date").reset_index(drop=True)
    s3 = s3.loc[s3["Date"] <= STUDY_END].sort_values(["Date", "Age_group"]).reset_index(drop=True)

    # Validate the broad age labels used by S3.
    observed_s3_ages = set(s3["Age_group"].dropna().astype(str).str.strip().unique())
    expected_s3_ages = {"All ages", "<60", "60+"}
    if observed_s3_ages != expected_s3_ages:
        raise ValueError(
            f"Unexpected S3 Age_group labels: {sorted(observed_s3_ages)}; "
            f"expected {sorted(expected_s3_ages)}"
        )

    return paths, s1, s2, s3, s4, s5


# =============================================================================
# 3. National epidemic trajectory (S1) and test positivity (S2)
# =============================================================================

def locate_wave_peaks(s1: pd.DataFrame) -> pd.DataFrame:
    """Locate the maximum 7-day moving average within each named wave window."""
    rows = []
    for wave, (start, end) in WAVE_WINDOWS.items():
        window = s1.loc[s1["Date"].between(pd.Timestamp(start), pd.Timestamp(end))]
        peak = window.loc[window["7_day_moving_average"].idxmax()]
        rows.append(
            {
                "wave": wave,
                "window_start": pd.Timestamp(start),
                "window_end": pd.Timestamp(end),
                "peak_date": peak["Date"],
                "peak_7dma_cases": float(peak["7_day_moving_average"]),
            }
        )
    return pd.DataFrame(rows)


def first_day_at_or_below_fraction(
    s1: pd.DataFrame,
    peak_date: pd.Timestamp,
    fraction: float,
) -> tuple[int | float, pd.Timestamp, float]:
    """Days from a peak to the first later day at/below a fraction of that peak."""
    peak_value = float(
        s1.loc[s1["Date"].eq(peak_date), "7_day_moving_average"].iloc[0]
    )
    threshold = peak_value * fraction
    eligible = s1.loc[
        (s1["Date"] > peak_date)
        & s1["7_day_moving_average"].notna()
        & (s1["7_day_moving_average"] <= threshold)
    ]
    if eligible.empty:
        return np.nan, pd.NaT, np.nan
    row = eligible.iloc[0]
    return (
        int((row["Date"] - peak_date).days),
        row["Date"],
        float(row["7_day_moving_average"]),
    )


def calculate_wave_resolution(s1: pd.DataFrame, wave_peaks: pd.DataFrame) -> pd.DataFrame:
    """Resolution metrics for the two pre-rollout waves and rollout-spanning wave."""
    target_waves = [
        "First pre-rollout wave",
        "Summer-autumn 2020 wave",
        "Winter 2020-2021 wave",
    ]
    rows = []
    for wave in target_waves:
        peak = wave_peaks.loc[wave_peaks["wave"].eq(wave)].iloc[0]
        result = {
            "wave": wave,
            "peak_date": peak["peak_date"],
            "peak_7dma_cases": peak["peak_7dma_cases"],
        }
        for fraction, label in [(0.50, "50pct"), (0.25, "25pct"), (0.10, "10pct")]:
            days, date_reached, value_reached = first_day_at_or_below_fraction(
                s1, peak["peak_date"], fraction
            )
            result[f"days_to_{label}"] = days
            result[f"date_at_{label}"] = date_reached
            result[f"value_at_{label}"] = value_reached
        rows.append(result)
    return pd.DataFrame(rows)


def calculate_test_positivity(s2: pd.DataFrame) -> pd.DataFrame:
    """Add a trailing 7-day moving average of overall test positivity."""
    out = s2.copy()
    out["Overall_positivity_7dma"] = (
        out["Overall_positivity_percent"].rolling(window=7, min_periods=1).mean()
    )
    return out


# =============================================================================
# 4. S3: routine-surveillance apparent vaccine effectiveness
# =============================================================================

def build_s3_weekly(
    s3: pd.DataFrame,
    age_group: str,
    min_cases_for_denominator: int = 1,
) -> pd.DataFrame:
    """
    Reconstruct weekly documented-infection rates from Dataset S3.

    Daily population denominator within each vaccination-status category:

        N = C * 100,000 / R

    where C is the reported daily number of cases and R is the reported rate
    per 100,000. In the primary analysis, denominators are reconstructed only
    when C>0 and R>0. For sensitivity analyses, C must instead be >=5 or >=10.

    The weekly denominator is the median of eligible daily reconstructed
    denominators; weekly cases are the sum of all daily cases.
    """
    g = s3.loc[s3["Age_group"].eq(age_group)].copy()
    g["week_start"] = sunday_week_start(g["Date"])

    categories = [
        ("Cases_vaccinated", "Cases_per_100k_vaccinated", "denom_vaccinated_daily"),
        (
            "Cases_expired_vaccination_status",
            "Cases_per_100k_expired_vaccination_status",
            "denom_expired_daily",
        ),
        ("Cases_unvaccinated", "Cases_per_100k_unvaccinated", "denom_unvaccinated_daily"),
    ]

    for cases_col, rate_col, denom_col in categories:
        eligible = (g[cases_col] >= min_cases_for_denominator) & (g[rate_col] > 0)
        g[denom_col] = np.where(
            eligible,
            g[cases_col] * 100000.0 / g[rate_col],
            np.nan,
        )

    weekly = (
        g.groupby("week_start", as_index=False)
        .agg(
            cases_vaccinated=("Cases_vaccinated", "sum"),
            cases_expired=("Cases_expired_vaccination_status", "sum"),
            cases_unvaccinated=("Cases_unvaccinated", "sum"),
            denominator_vaccinated=("denom_vaccinated_daily", "median"),
            denominator_expired=("denom_expired_daily", "median"),
            denominator_unvaccinated=("denom_unvaccinated_daily", "median"),
        )
        .sort_values("week_start")
        .reset_index(drop=True)
    )

    # Combine IMOH 'vaccinated' and 'vaccination status expired' categories.
    weekly["cases_combined_vaccinated"] = weekly[
        ["cases_vaccinated", "cases_expired"]
    ].sum(axis=1, min_count=1)
    weekly["denominator_combined_vaccinated"] = weekly[
        ["denominator_vaccinated", "denominator_expired"]
    ].sum(axis=1, min_count=1)

    weekly["rate_per_100k_combined_vaccinated"] = (
        weekly["cases_combined_vaccinated"]
        / weekly["denominator_combined_vaccinated"]
        * 100000.0
    )
    weekly["rate_per_100k_unvaccinated"] = (
        weekly["cases_unvaccinated"]
        / weekly["denominator_unvaccinated"]
        * 100000.0
    )
    weekly["RR_cases"] = (
        weekly["rate_per_100k_combined_vaccinated"]
        / weekly["rate_per_100k_unvaccinated"]
    )
    weekly["VE_surv_percent"] = 100.0 * (1.0 - weekly["RR_cases"])
    weekly["Age_group"] = age_group
    weekly["min_cases_for_denominator"] = min_cases_for_denominator

    return weekly


# =============================================================================
# 5. S4-S5: harmonization, consistency check, and linked analysis cells
# =============================================================================

def harmonize_s5_age(value: str) -> str:
    """Map S5 age categories onto the eight age categories used by S4."""
    if value in {"0-9", "10-19"}:
        return "0-19"
    if value in {"80-89", "90+"}:
        return "80+"
    return value


def prepare_s4_s5(
    s4_raw: pd.DataFrame,
    s5_raw: pd.DataFrame,
    suppressed_value: float = SUPPRESSED_PRIMARY,
):
    """
    Prepare S4-S5 for the testing/prior-infection-informed analysis.

    Privacy-suppressed '<5' cells are assigned the requested value. S5 is
    harmonized to S4's eight age groups. The primary analysis then selects
    vaccinated and unvaccinated people without a previous documented infection.
    """
    s4 = s4_raw.copy()
    s5 = s5_raw.copy()

    s4_count_cols = [c for c in s4.columns if c not in {"Week", "Age_group"}]
    s5_count_cols = [c for c in s5.columns if c not in {"Test_week", "Age_group"}]

    for col in s4_count_cols:
        s4[col] = parse_suppressed_count(s4[col], suppressed_value)
    for col in s5_count_cols:
        s5[col] = parse_suppressed_count(s5[col], suppressed_value)

    s4["week_start"] = parse_week_start(s4["Week"])
    s5["week_start"] = parse_week_start(s5["Test_week"])

    s4["Age_group_harmonized"] = s4["Age_group"].astype(str).str.strip()
    s5["Age_group_harmonized"] = (
        s5["Age_group"].astype(str).str.strip().map(harmonize_s5_age)
    )

    # Common analytical period of S4 and S5.
    s4 = s4.loc[s4["week_start"].between(LINKED_START, LINKED_LAST_WEEK)].copy()
    s5 = s5.loc[s5["week_start"].between(LINKED_START, LINKED_LAST_WEEK)].copy()

    # S4 already has the target eight age groups; aggregate S5 to those groups.
    s4h = (
        s4.groupby(["week_start", "Age_group_harmonized"], as_index=False)[s4_count_cols]
        .sum(min_count=1)
    )
    s5h = (
        s5.groupby(["week_start", "Age_group_harmonized"], as_index=False)[s5_count_cols]
        .sum(min_count=1)
    )

    for df in (s4h, s5h):
        df["Age_group_harmonized"] = pd.Categorical(
            df["Age_group_harmonized"], categories=AGE_ORDER, ordered=True
        )
        df.sort_values(["week_start", "Age_group_harmonized"], inplace=True)
        df.reset_index(drop=True, inplace=True)

    # -------------------------------------------------------------------------
    # 5a. Internal consistency check
    # -------------------------------------------------------------------------
    # Compare S4 Tests_num with the total reconstructed by summing all S5
    # vaccination/recovery-specific testing categories.
    consistency = s4h[["week_start", "Age_group_harmonized", "Tests_num"]].merge(
        s5h[["week_start", "Age_group_harmonized"] + s5_count_cols],
        on=["week_start", "Age_group_harmonized"],
        how="inner",
        validate="one_to_one",
    )
    consistency["S5_reconstructed_total_tests"] = consistency[s5_count_cols].sum(axis=1)
    consistency["S5_to_S4_ratio"] = (
        consistency["S5_reconstructed_total_tests"] / consistency["Tests_num"]
    )
    consistency["absolute_relative_difference"] = (
        (consistency["S5_reconstructed_total_tests"] - consistency["Tests_num"]).abs()
        / consistency["Tests_num"]
    )

    # -------------------------------------------------------------------------
    # 5b. Primary vaccinated-unvaccinated comparison
    # -------------------------------------------------------------------------
    linked = s4h[
        [
            "week_start", "Age_group_harmonized",
            "Not_recovered_at_least_2_doses",
            "Not_recovered_not_vaccinated",
        ]
    ].merge(
        s5h[
            [
                "week_start", "Age_group_harmonized",
                "Non_recovered_not_vaccinated",
                "Non_recovered_2_doses",
                "Non_recovered_3_and_more_doses",
            ]
        ],
        on=["week_start", "Age_group_harmonized"],
        how="inner",
        validate="one_to_one",
    )

    linked["vaccinated_cases"] = linked["Not_recovered_at_least_2_doses"]
    linked["unvaccinated_cases"] = linked["Not_recovered_not_vaccinated"]
    linked["vaccinated_tests"] = (
        linked["Non_recovered_2_doses"]
        + linked["Non_recovered_3_and_more_doses"]
    )
    linked["unvaccinated_tests"] = linked["Non_recovered_not_vaccinated"]

    linked["vaccinated_positivity"] = (
        linked["vaccinated_cases"] / linked["vaccinated_tests"]
    )
    linked["unvaccinated_positivity"] = (
        linked["unvaccinated_cases"] / linked["unvaccinated_tests"]
    )

    return s4h, s5h, consistency, linked


# =============================================================================
# 6. Mantel-Haenszel risk ratio and Greenland-Robins 95% CI
# =============================================================================

def mantel_haenszel_rr_greenland_robins(strata: pd.DataFrame) -> pd.Series:
    """
    Mantel-Haenszel common risk ratio with Greenland-Robins log-RR variance.

    For each stratum h:
        a_h  = vaccinated cases
        c_h  = unvaccinated cases
        n1_h = vaccinated tested
        n0_h = unvaccinated tested
        n_h  = n1_h + n0_h

        RR_MH = sum(a_h*n0_h/n_h) / sum(c_h*n1_h/n_h)

    Greenland-Robins variance of log(RR_MH):

        sum([n1_h*n0_h*(a_h+c_h) - a_h*c_h*n_h] / n_h^2) / (A*B)

    where A and B are the numerator and denominator sums in RR_MH.
    """
    x = strata.loc[
        (strata["vaccinated_tests"] > 0) & (strata["unvaccinated_tests"] > 0)
    ].copy()

    a = x["vaccinated_cases"].astype(float)
    c = x["unvaccinated_cases"].astype(float)
    n1 = x["vaccinated_tests"].astype(float)
    n0 = x["unvaccinated_tests"].astype(float)
    n = n1 + n0

    A = (a * n0 / n).sum()
    B = (c * n1 / n).sum()

    if A <= 0 or B <= 0:
        return pd.Series(
            {
                "RR_MH": np.nan,
                "RR_low": np.nan,
                "RR_high": np.nan,
                "SE_log_RR": np.nan,
                "VE_percent": np.nan,
                "VE_low_percent": np.nan,
                "VE_high_percent": np.nan,
                "n_strata": len(x),
            }
        )

    rr = A / B
    variance_numerator = (
        (n1 * n0 * (a + c) - a * c * n) / (n ** 2)
    ).sum()
    var_log_rr = variance_numerator / (A * B)
    se_log_rr = math.sqrt(var_log_rr)

    rr_low = rr * math.exp(-Z_975 * se_log_rr)
    rr_high = rr * math.exp(Z_975 * se_log_rr)

    return pd.Series(
        {
            "RR_MH": rr,
            "RR_low": rr_low,
            "RR_high": rr_high,
            "SE_log_RR": se_log_rr,
            "VE_percent": 100.0 * (1.0 - rr),
            # RR confidence limits reverse order after transformation to VE.
            "VE_low_percent": 100.0 * (1.0 - rr_high),
            "VE_high_percent": 100.0 * (1.0 - rr_low),
            "n_strata": len(x),
        }
    )


def weekly_mh(linked: pd.DataFrame, ages: list[str], label: str) -> pd.DataFrame:
    """Calculate one age-adjusted Mantel-Haenszel estimate per calendar week."""
    x = linked.loc[linked["Age_group_harmonized"].astype(str).isin(ages)].copy()
    rows = []
    for week_start, week_data in x.groupby("week_start", sort=True):
        est = mantel_haenszel_rr_greenland_robins(week_data)
        rows.append(
            {"week_start": week_start, "analysis_age_group": label, **est.to_dict()}
        )
    return pd.DataFrame(rows).sort_values("week_start").reset_index(drop=True)


def full_period_mh(linked: pd.DataFrame, ages: list[str], label: str) -> pd.Series:
    """
    Full-period sensitivity summary with every age-by-week row as a stratum.
    """
    x = linked.loc[linked["Age_group_harmonized"].astype(str).isin(ages)].copy()
    est = mantel_haenszel_rr_greenland_robins(x)
    est["analysis"] = label
    return est


# =============================================================================
# 7. Compare trajectories and run sensitivity analyses
# =============================================================================

def compare_primary_trajectories(
    s3_all: pd.DataFrame,
    mh_all: pd.DataFrame,
) -> tuple[pd.DataFrame, dict]:
    """Directly compare the two all-age weekly trajectories."""
    comparison = s3_all[["week_start", "RR_cases", "VE_surv_percent"]].merge(
        mh_all[
            [
                "week_start", "RR_MH", "RR_low", "RR_high",
                "VE_percent", "VE_low_percent", "VE_high_percent",
            ]
        ],
        on="week_start",
        how="inner",
        validate="one_to_one",
    )
    comparison = comparison.loc[
        comparison["week_start"].between(LINKED_START, LINKED_LAST_WEEK)
    ].copy()
    comparison.rename(
        columns={"VE_percent": "VE_testing_informed_percent"}, inplace=True
    )

    routine_positive = comparison.loc[comparison["VE_surv_percent"] > 0].copy()
    summary = {
        "overlapping_weeks": int(len(comparison)),
        "routine_positive_weeks": int(len(routine_positive)),
        "testing_lower_when_routine_positive": int(
            (routine_positive["VE_testing_informed_percent"] < routine_positive["VE_surv_percent"]).sum()
        ),
        "testing_negative_when_routine_positive": int(
            (routine_positive["VE_testing_informed_percent"] < 0).sum()
        ),
        "median_routine_among_routine_positive": float(
            routine_positive["VE_surv_percent"].median()
        ),
        "median_testing_among_routine_positive": float(
            routine_positive["VE_testing_informed_percent"].median()
        ),
        "routine_negative_testing_positive_weeks": int(
            (
                (comparison["VE_surv_percent"] < 0)
                & (comparison["VE_testing_informed_percent"] > 0)
            ).sum()
        ),
    }
    return comparison, summary


def suppressed_cell_sensitivity(
    s4_raw: pd.DataFrame,
    s5_raw: pd.DataFrame,
    mh_primary: pd.DataFrame,
) -> tuple[pd.DataFrame, dict[float, pd.DataFrame]]:
    """Repeat weekly S4-S5 estimates assigning '<5' cells 1 and 4."""
    rows = []
    weekly_outputs = {}
    for value in [1.0, 4.0]:
        _, _, _, linked_alt = prepare_s4_s5(
            s4_raw, s5_raw, suppressed_value=value
        )
        mh_alt = weekly_mh(linked_alt, AGE_ORDER, f"All ages; <5={value:g}")
        weekly_outputs[value] = mh_alt

        merged = mh_primary[["week_start", "VE_percent"]].merge(
            mh_alt[["week_start", "VE_percent"]],
            on="week_start",
            how="inner",
            suffixes=("_primary", "_alternative"),
        ).dropna()
        absolute_difference = (
            merged["VE_percent_alternative"] - merged["VE_percent_primary"]
        ).abs()
        rows.append(
            {
                "suppressed_cell_assignment": value,
                "n_comparable_weeks": len(merged),
                "median_absolute_difference_percentage_points": float(
                    absolute_difference.median()
                ),
                "maximum_absolute_difference_percentage_points": float(
                    absolute_difference.max()
                ),
            }
        )

    return pd.DataFrame(rows), weekly_outputs


def s3_denominator_sensitivity(
    s3: pd.DataFrame,
    s3_primary: pd.DataFrame,
) -> tuple[pd.DataFrame, dict[int, pd.DataFrame]]:
    """Repeat S3 reconstruction using minimum daily case counts of 5 and 10."""
    rows = []
    weekly_outputs = {}
    for threshold in [5, 10]:
        alternative = build_s3_weekly(
            s3, "All ages", min_cases_for_denominator=threshold
        )
        weekly_outputs[threshold] = alternative

        merged = s3_primary[["week_start", "VE_surv_percent"]].merge(
            alternative[["week_start", "VE_surv_percent"]],
            on="week_start",
            how="inner",
            suffixes=("_primary", f"_min{threshold}"),
        ).dropna()
        absolute_difference = (
            merged[f"VE_surv_percent_min{threshold}"]
            - merged["VE_surv_percent_primary"]
        ).abs()
        rows.append(
            {
                "minimum_daily_cases_for_denominator": threshold,
                "n_comparable_weeks": len(merged),
                "median_absolute_difference_percentage_points": float(
                    absolute_difference.median()
                ),
                "maximum_absolute_difference_percentage_points": float(
                    absolute_difference.max()
                ),
            }
        )

    return pd.DataFrame(rows), weekly_outputs


# =============================================================================
# 8. Verify the exact manuscript-facing numerical results
# =============================================================================

def verify_manuscript_results(
    s1: pd.DataFrame,
    wave_peaks: pd.DataFrame,
    wave_resolution: pd.DataFrame,
    consistency: pd.DataFrame,
    s3_all: pd.DataFrame,
    mh_all: pd.DataFrame,
    comparison_summary: dict,
    suppression_sensitivity: pd.DataFrame,
    s3_sensitivity: pd.DataFrame,
    full_primary: pd.Series,
    full_adult: pd.Series,
) -> None:
    """
    Fail loudly if the supplied five datasets do not reproduce the numerical
    results reported in the current manuscript.
    """

    # 8a. National epidemic trajectory
    expected_peaks = {
        "First pre-rollout wave": ("2020-04-06", 616.0),
        "Summer-autumn 2020 wave": ("2020-09-28", 6283.0),
        "Winter 2020-2021 wave": ("2021-01-16", 8333.0),
        "Delta-period wave": ("2021-09-04", 9746.0),
        "Omicron BA.1-period wave": ("2022-01-26", 75624.0),
        "Spring 2022 wave": ("2022-03-30", 13321.0),
        "Summer 2022 wave": ("2022-07-05", 10861.0),
    }
    for wave, (expected_date, expected_peak) in expected_peaks.items():
        row = wave_peaks.loc[wave_peaks["wave"].eq(wave)].iloc[0]
        if fmt_date(row["peak_date"]) != expected_date:
            raise AssertionError(
                f"{wave}: peak date {fmt_date(row['peak_date'])} != {expected_date}"
            )
        assert_close(f"{wave} peak", row["peak_7dma_cases"], expected_peak)

    expected_resolution = {
        "First pre-rollout wave": (15, 25, 32),
        "Summer-autumn 2020 wave": (15, 22, 43),
        "Winter 2020-2021 wave": (33, 60, 68),
    }
    for wave, expected in expected_resolution.items():
        row = wave_resolution.loc[wave_resolution["wave"].eq(wave)].iloc[0]
        observed = (
            int(row["days_to_50pct"]),
            int(row["days_to_25pct"]),
            int(row["days_to_10pct"]),
        )
        if observed != expected:
            raise AssertionError(f"{wave}: resolution {observed} != {expected}")

    rollout_value = float(
        s1.loc[s1["Date"].eq(ROLLOUT_START), "7_day_moving_average"].iloc[0]
    )
    assert_close("7-day moving average at vaccine rollout", rollout_value, 2475.0)

    # 8b. S4-S5 consistency
    if len(consistency) != 832:
        raise AssertionError(f"Expected 832 matched week-by-age cells; found {len(consistency)}")
    assert_close(
        "median S5/S4 ratio",
        consistency["S5_to_S4_ratio"].median(),
        0.9880522740630485,
    )
    assert_close(
        "median absolute relative S5/S4 difference",
        consistency["absolute_relative_difference"].median(),
        0.011947725936951516,
    )
    if int((consistency["absolute_relative_difference"] <= 0.05).sum()) != 806:
        raise AssertionError("Expected 806 of 832 cells to differ by no more than 5%")

    # Verify the different starting dates of the two vaccination-status analyses.
    s3_first_week = pd.Timestamp(s3_all["week_start"].min())
    mh_first_week = pd.Timestamp(mh_all["week_start"].min())
    if s3_first_week != pd.Timestamp("2021-01-17"):
        raise AssertionError(
            f"Expected S3 routine-surveillance analysis to begin 2021-01-17; "
            f"found {fmt_date(s3_first_week)}"
        )
    if mh_first_week != LINKED_START:
        raise AssertionError(
            f"Expected S4-S5 informed analysis to begin {fmt_date(LINKED_START)}; "
            f"found {fmt_date(mh_first_week)}"
        )
    if (mh_first_week - s3_first_week).days != 19 * 7:
        raise AssertionError(
            "Expected the S4-S5 informed analysis to begin exactly 19 weeks "
            "after the S3 routine-surveillance analysis"
        )

    # 8c. S3 routine-surveillance values
    expected_s3 = {
        "2021-02-14": 94.40513489684359,
        "2021-05-30": 61.46179401993356,
        "2021-09-12": 72.31506921487670,
        "2021-12-05": 79.89593387748217,
        "2021-12-19": 37.41554520728019,
        "2021-12-26": 8.18522502613106,
        "2022-01-02": -3.71416880465791,
        "2022-02-13": -3.34117354245189,
    }
    for week, expected in expected_s3.items():
        assert_close(
            f"S3 VE {week}",
            get_week_row(s3_all, week)["VE_surv_percent"],
            expected,
            atol=1e-7,
        )

    if not (
        s3_all.loc[s3_all["week_start"] >= pd.Timestamp("2022-02-13"), "VE_surv_percent"] < 0
    ).all():
        raise AssertionError("S3 estimate should remain negative from 2022-02-13 onward")

    # 8d. S4-S5 testing/prior-infection-informed values
    expected_mh = {
        "2021-05-30": (0.25039539926721716, 0.15148456507232222, 0.41388940149945774),
        "2021-09-12": (0.5842300268064479, 0.570885, 0.597887),
        "2021-12-05": (0.3785990970654295, 0.345045, 0.415416),
        "2021-12-19": (1.1453778667370942, 1.0864294825372665, 1.2075247208382127),
    }
    for week, (rr, lo, hi) in expected_mh.items():
        row = get_week_row(mh_all, week)
        assert_close(f"MH RR {week}", row["RR_MH"], rr, atol=1e-8)
        assert_close(f"MH RR lower CI {week}", row["RR_low"], lo, atol=2e-6)
        assert_close(f"MH RR upper CI {week}", row["RR_high"], hi, atol=2e-6)

    if not (
        mh_all.loc[mh_all["week_start"] >= pd.Timestamp("2021-12-19"), "VE_percent"] < 0
    ).all():
        raise AssertionError(
            "Testing/prior-infection-informed estimate should remain negative "
            "from 2021-12-19 onward"
        )

    # 8e. Direct comparison
    expected_comparison = {
        "overlapping_weeks": 104,
        "routine_positive_weeks": 36,
        "testing_lower_when_routine_positive": 35,
        "testing_negative_when_routine_positive": 14,
        "routine_negative_testing_positive_weeks": 0,
    }
    for key, expected in expected_comparison.items():
        if comparison_summary[key] != expected:
            raise AssertionError(
                f"{key}: observed={comparison_summary[key]}, expected={expected}"
            )
    assert_close(
        "median routine VE among routine-positive weeks",
        comparison_summary["median_routine_among_routine_positive"],
        55.109828238178196,
        atol=1e-8,
    )
    assert_close(
        "median testing-informed VE among routine-positive weeks",
        comparison_summary["median_testing_among_routine_positive"],
        17.628177148884166,
        atol=1e-8,
    )

    # 8f. Sensitivity analyses
    for _, row in suppression_sensitivity.iterrows():
        assert_close(
            f"<5 sensitivity median difference, assignment={row['suppressed_cell_assignment']}",
            row["median_absolute_difference_percentage_points"],
            0.0,
        )
    for _, row in s3_sensitivity.iterrows():
        assert_close(
            f"S3 denominator sensitivity median difference, threshold={row['minimum_daily_cases_for_denominator']}",
            row["median_absolute_difference_percentage_points"],
            0.0,
        )
    assert_close(
        "full-period primary VE-scale summary",
        full_primary["VE_percent"],
        -36.40966968276829,
        atol=1e-8,
    )
    assert_close(
        "full-period adults >=20 VE-scale summary",
        full_adult["VE_percent"],
        -41.87907735866403,
        atol=1e-8,
    )


# =============================================================================
# 9. Export a concise manuscript-facing result summary
# =============================================================================

def write_results_summary(
    output_dir: Path,
    s1: pd.DataFrame,
    positivity: pd.DataFrame,
    wave_peaks: pd.DataFrame,
    wave_resolution: pd.DataFrame,
    consistency: pd.DataFrame,
    s3_all: pd.DataFrame,
    mh_all: pd.DataFrame,
    comparison_summary: dict,
    suppression_sensitivity: pd.DataFrame,
    s3_sensitivity: pd.DataFrame,
    full_primary: pd.Series,
    full_adult: pd.Series,
) -> None:
    lines: list[str] = []
    add = lines.append

    add("MANUSCRIPT-FACING REPRODUCIBILITY SUMMARY")
    add("=========================================")
    add("")

    add("National epidemic trajectory")
    for _, row in wave_peaks.iterrows():
        add(
            f"- {row['wave']}: peak {row['peak_7dma_cases']:.0f} "
            f"on {fmt_date(row['peak_date'])}"
        )
    for _, row in wave_resolution.iterrows():
        add(
            f"- {row['wave']} resolution: <=50% in {int(row['days_to_50pct'])} d; "
            f"<=25% in {int(row['days_to_25pct'])} d; "
            f"<=10% in {int(row['days_to_10pct'])} d"
        )
    rollout = float(
        s1.loc[s1["Date"].eq(ROLLOUT_START), "7_day_moving_average"].iloc[0]
    )
    winter_peak = float(
        wave_peaks.loc[
            wave_peaks["wave"].eq("Winter 2020-2021 wave"), "peak_7dma_cases"
        ].iloc[0]
    )
    add(
        f"- At vaccine rollout (2020-12-20): 7dma={rollout:.0f}, "
        f"{100 * rollout / winter_peak:.1f}% of the eventual winter peak"
    )
    maximum_positivity = positivity.loc[positivity["Overall_positivity_7dma"].idxmax()]
    add(
        f"- Maximum 7-day overall test positivity: "
        f"{maximum_positivity['Overall_positivity_7dma']:.1f}% "
        f"on {fmt_date(maximum_positivity['Date'])}"
    )
    add("")

    add("S4-S5 consistency")
    add(f"- Matched week-by-age cells: {len(consistency)}")
    add(f"- Median S5/S4 ratio: {consistency['S5_to_S4_ratio'].median():.3f}")
    add(
        f"- Median absolute relative difference: "
        f"{100 * consistency['absolute_relative_difference'].median():.2f}%"
    )
    within_5 = int((consistency["absolute_relative_difference"] <= 0.05).sum())
    add(f"- Within 5%: {within_5}/{len(consistency)} ({100 * within_5 / len(consistency):.1f}%)")
    add("")

    add("Routine-surveillance apparent VE (S3)")
    for week in [
        "2021-02-14", "2021-05-30", "2021-09-12", "2021-12-05",
        "2021-12-19", "2021-12-26", "2022-01-02", "2022-02-13",
    ]:
        row = get_week_row(s3_all, week)
        add(f"- {week}: {row['VE_surv_percent']:.1f}%")
    add("")

    add("Testing- and prior-infection-informed estimate")
    for week in ["2021-05-30", "2021-09-12", "2021-12-05", "2021-12-19"]:
        row = get_week_row(mh_all, week)
        add(
            f"- {week}: RR={row['RR_MH']:.3f} "
            f"(95% CI {row['RR_low']:.3f}-{row['RR_high']:.3f}); "
            f"VE-scale={row['VE_percent']:.1f}% "
            f"(95% CI {row['VE_low_percent']:.1f}% to {row['VE_high_percent']:.1f}%)"
        )
    add("- Negative in every week from 2021-12-19 through the end of follow-up.")
    add("")

    add("Direct comparison")
    add(f"- Overlapping weeks: {comparison_summary['overlapping_weeks']}")
    add(f"- Routine-surveillance estimate positive: {comparison_summary['routine_positive_weeks']} weeks")
    add(
        f"- Among those weeks, testing-informed estimate was lower in "
        f"{comparison_summary['testing_lower_when_routine_positive']} and already negative in "
        f"{comparison_summary['testing_negative_when_routine_positive']}"
    )
    add(
        f"- Median estimates in those weeks: routine="
        f"{comparison_summary['median_routine_among_routine_positive']:.1f}%; "
        f"testing-informed={comparison_summary['median_testing_among_routine_positive']:.1f}%"
    )
    add(
        f"- Weeks with routine estimate negative but testing-informed estimate positive: "
        f"{comparison_summary['routine_negative_testing_positive_weeks']}"
    )
    add("")

    add("Sensitivity analyses")
    for _, row in suppression_sensitivity.iterrows():
        add(
            f"- <5 assigned {row['suppressed_cell_assignment']:.0f}: "
            f"median weekly absolute difference="
            f"{row['median_absolute_difference_percentage_points']:.1f} percentage points"
        )
    add(
        f"- Full-period age-by-week stratified VE-scale summary: "
        f"primary={full_primary['VE_percent']:.1f}%; adults >=20={full_adult['VE_percent']:.1f}%"
    )
    for _, row in s3_sensitivity.iterrows():
        add(
            f"- S3 denominator reconstruction threshold >="
            f"{int(row['minimum_daily_cases_for_denominator'])} cases: "
            f"median weekly absolute difference="
            f"{row['median_absolute_difference_percentage_points']:.1f} percentage points"
        )

    text = "\n".join(lines) + "\n"
    (output_dir / "manuscript_results_summary.txt").write_text(text, encoding="utf-8")
    print(text)


# =============================================================================
# 10. Optional figures
# =============================================================================

def make_figures(
    s1: pd.DataFrame,
    positivity: pd.DataFrame,
    wave_peaks: pd.DataFrame,
    s3_all: pd.DataFrame,
    mh_all: pd.DataFrame,
    output_dir: Path,
) -> None:
    """Generate reproducible versions of Figures 1-3 from the analysis data."""
    figure_dir = output_dir / "figures"
    figure_dir.mkdir(parents=True, exist_ok=True)

    # Figure 1: full national case trajectory plus expanded early period.
    fig, axes = plt.subplots(2, 1, figsize=(11, 8), constrained_layout=True)

    axes[0].plot(s1["Date"], s1["7_day_moving_average"], linewidth=1.5)
    axes[0].set_title("A. Full national trajectory")
    axes[0].set_ylabel("Daily confirmed cases\n(7-day moving average)")
    axes[0].set_xlim(s1["Date"].min(), STUDY_END)

    early = s1.loc[s1["Date"].between(pd.Timestamp("2020-02-12"), pd.Timestamp("2021-05-31"))]
    axes[1].plot(early["Date"], early["7_day_moving_average"], linewidth=1.5)
    axes[1].axvline(ROLLOUT_START, linestyle="--", linewidth=1.0)
    axes[1].set_title("B. Expanded pre-rollout and initial-rollout period")
    axes[1].set_ylabel("Daily confirmed cases\n(7-day moving average)")
    axes[1].set_xlabel("Date")

    for wave in [
        "First pre-rollout wave",
        "Summer-autumn 2020 wave",
        "Winter 2020-2021 wave",
    ]:
        row = wave_peaks.loc[wave_peaks["wave"].eq(wave)].iloc[0]
        axes[1].annotate(
            f"{row['peak_date'].strftime('%b %d')}\n{row['peak_7dma_cases']:.0f}",
            xy=(row["peak_date"], row["peak_7dma_cases"]),
            xytext=(0, 8),
            textcoords="offset points",
            ha="center",
            fontsize=8,
        )

    fig.suptitle("National trajectory of documented SARS-CoV-2 infection in Israel")
    fig.savefig(figure_dir / "Figure1_National_Cases.png", dpi=300, bbox_inches="tight")
    plt.close(fig)

    # Figure 2: national test positivity.
    fig, ax = plt.subplots(figsize=(11, 5))
    ax.plot(positivity["Date"], positivity["Overall_positivity_7dma"], linewidth=1.5)
    ax.set_title("National trajectory of SARS-CoV-2 test positivity")
    ax.set_ylabel("Overall test positivity (%)\n7-day moving average")
    ax.set_xlabel("Date")
    ax.set_xlim(positivity["Date"].min(), STUDY_END)
    fig.tight_layout()
    fig.savefig(figure_dir / "Figure2_Test_Positivity.png", dpi=300, bbox_inches="tight")
    plt.close(fig)

    # Figure 3: retain the full S3 routine-surveillance trajectory, including
    # the 19 weeks before the S4-S5 testing/prior-infection-informed analysis
    # becomes available.
    fig, ax = plt.subplots(figsize=(11.5, 5.8))
    ax.plot(
        s3_all["week_start"],
        s3_all["VE_surv_percent"],
        label="Routine surveillance (S3)",
        linewidth=1.6,
    )
    ax.plot(
        mh_all["week_start"],
        mh_all["VE_percent"],
        label="Testing- and prior-infection-informed (S4-S5)",
        linewidth=1.6,
    )
    ax.axhline(0, linewidth=0.9)
    ax.axvline(LINKED_START, linestyle="--", linewidth=1.0)
    ax.annotate(
        "Start of testing- and prior-infection-informed analysis\n"
        "(week beginning May 30, 2021)",
        xy=(LINKED_START, 61.5),
        xytext=(20, 18),
        textcoords="offset points",
        fontsize=8.5,
    )
    ax.set_title("Overall vaccinated-unvaccinated contrast over time")
    ax.set_ylabel("VE-scale contrast (%)")
    ax.set_xlabel("Week beginning")
    ax.set_xlim(pd.Timestamp("2021-01-17"), STUDY_END)
    ax.xaxis.set_major_locator(mdates.MonthLocator(interval=3))
    ax.xaxis.set_major_formatter(mdates.DateFormatter("%b\n%Y"))
    ax.legend(frameon=False)
    fig.tight_layout()

    fig.savefig(
        figure_dir / "Figure3_Overall_Comparison.png",
        dpi=300,
        bbox_inches="tight",
    )
    fig.savefig(
        figure_dir / "Figure3_Overall_Comparison.svg",
        bbox_inches="tight",
    )
    plt.close(fig)


# =============================================================================
# 11. Main reproducibility workflow
# =============================================================================

def main() -> None:
    parser = argparse.ArgumentParser(
        description="Reproduce the Israel national COVID-19 surveillance analyses."
    )
    parser.add_argument(
        "--data-dir",
        type=Path,
        default=Path(__file__).resolve().parent,
        help="Folder containing Supplementary Datasets S1-S5.",
    )
    parser.add_argument(
        "--output-dir",
        type=Path,
        default=None,
        help="Output folder (default: <data-dir>/analysis_outputs).",
    )
    parser.add_argument(
        "--no-figures",
        action="store_true",
        help="Run all analyses but do not generate figure files.",
    )
    args = parser.parse_args()

    data_dir = args.data_dir.resolve()
    output_dir = (args.output_dir or (data_dir / "analysis_outputs")).resolve()
    tables_dir = output_dir / "tables"
    output_dir.mkdir(parents=True, exist_ok=True)
    tables_dir.mkdir(parents=True, exist_ok=True)

    # -------------------------------------------------------------------------
    # Step 1. Load the five datasets
    # -------------------------------------------------------------------------
    paths, s1, s2, s3, s4_raw, s5_raw = load_datasets(data_dir)

    print("Input files and SHA-256 checksums:")
    provenance_rows = []
    for key, path in paths.items():
        digest = sha256(path)
        print(f"  {key}: {path.name}  {digest}")
        provenance_rows.append(
            {"dataset": key, "filename": path.name, "sha256": digest}
        )
    pd.DataFrame(provenance_rows).to_csv(
        output_dir / "input_file_checksums.csv", index=False
    )

    # -------------------------------------------------------------------------
    # Step 2. National epidemic trajectory
    # -------------------------------------------------------------------------
    wave_peaks = locate_wave_peaks(s1)
    wave_resolution = calculate_wave_resolution(s1, wave_peaks)
    positivity = calculate_test_positivity(s2)

    # -------------------------------------------------------------------------
    # Step 3. Routine-surveillance trajectory from S3
    # -------------------------------------------------------------------------
    s3_all = build_s3_weekly(s3, "All ages", min_cases_for_denominator=1)
    s3_lt60 = build_s3_weekly(s3, "<60", min_cases_for_denominator=1)
    s3_60plus = build_s3_weekly(s3, "60+", min_cases_for_denominator=1)

    # -------------------------------------------------------------------------
    # Step 4. Harmonize/link S4-S5 and perform the internal consistency check
    # -------------------------------------------------------------------------
    s4_harmonized, s5_harmonized, consistency, linked = prepare_s4_s5(
        s4_raw, s5_raw, suppressed_value=SUPPRESSED_PRIMARY
    )

    # -------------------------------------------------------------------------
    # Step 5. Weekly age-adjusted testing/prior-infection-informed estimates
    # -------------------------------------------------------------------------
    mh_all = weekly_mh(linked, AGE_ORDER, "All ages")
    mh_lt60 = weekly_mh(linked, AGES_LT60, "<60")
    mh_60plus = weekly_mh(linked, AGES_60PLUS, "60+")

    # -------------------------------------------------------------------------
    # Step 6. Directly compare the two primary trajectories
    # -------------------------------------------------------------------------
    comparison, comparison_summary = compare_primary_trajectories(s3_all, mh_all)

    # -------------------------------------------------------------------------
    # Step 7. Sensitivity analyses
    # -------------------------------------------------------------------------
    suppression_sens, suppression_weekly = suppressed_cell_sensitivity(
        s4_raw, s5_raw, mh_all
    )
    s3_sens, s3_sensitivity_weekly = s3_denominator_sensitivity(s3, s3_all)

    full_primary = full_period_mh(
        linked, AGE_ORDER, "All ages; age-by-week strata"
    )
    full_adult = full_period_mh(
        linked, AGES_ADULT, "Adults >=20; age-by-week strata"
    )

    # -------------------------------------------------------------------------
    # Step 8. Verify that this run reproduces the manuscript Results exactly
    # -------------------------------------------------------------------------
    verify_manuscript_results(
        s1=s1,
        wave_peaks=wave_peaks,
        wave_resolution=wave_resolution,
        consistency=consistency,
        s3_all=s3_all,
        mh_all=mh_all,
        comparison_summary=comparison_summary,
        suppression_sensitivity=suppression_sens,
        s3_sensitivity=s3_sens,
        full_primary=full_primary,
        full_adult=full_adult,
    )

    # -------------------------------------------------------------------------
    # Step 9. Export transparent intermediate and final tables
    # -------------------------------------------------------------------------
    wave_peaks.to_csv(tables_dir / "01_wave_peaks.csv", index=False)
    wave_resolution.to_csv(tables_dir / "02_wave_resolution.csv", index=False)
    positivity[["Date", "Overall_positivity_percent", "Overall_positivity_7dma"]].to_csv(
        tables_dir / "03_national_test_positivity.csv", index=False
    )

    s4_harmonized.to_csv(tables_dir / "04_S4_harmonized.csv", index=False)
    s5_harmonized.to_csv(tables_dir / "05_S5_harmonized.csv", index=False)
    consistency.to_csv(tables_dir / "06_S4_S5_consistency_check.csv", index=False)
    linked.to_csv(tables_dir / "07_S4_S5_linked_analysis_cells.csv", index=False)

    pd.concat([s3_all, s3_lt60, s3_60plus], ignore_index=True).to_csv(
        tables_dir / "08_S3_weekly_apparent_surveillance_VE.csv", index=False
    )
    pd.concat([mh_all, mh_lt60, mh_60plus], ignore_index=True).to_csv(
        tables_dir / "09_S4_S5_weekly_testing_priorinfection_informed_VE.csv",
        index=False,
    )
    comparison.to_csv(
        tables_dir / "10_primary_trajectory_comparison.csv", index=False
    )

    suppression_sens.to_csv(
        tables_dir / "11_sensitivity_suppressed_cells_summary.csv", index=False
    )
    s3_sens.to_csv(
        tables_dir / "12_sensitivity_S3_denominator_summary.csv", index=False
    )
    pd.DataFrame([full_primary.to_dict(), full_adult.to_dict()]).to_csv(
        tables_dir / "13_full_period_age_week_stratified_sensitivity.csv", index=False
    )

    for value, table in suppression_weekly.items():
        table.to_csv(
            tables_dir / f"sensitivity_weekly_suppressed_cells_{int(value)}.csv",
            index=False,
        )
    for threshold, table in s3_sensitivity_weekly.items():
        table.to_csv(
            tables_dir / f"sensitivity_S3_min_cases_{threshold}.csv",
            index=False,
        )

    # -------------------------------------------------------------------------
    # Step 10. Write a concise result summary and optional figures
    # -------------------------------------------------------------------------
    write_results_summary(
        output_dir=output_dir,
        s1=s1,
        positivity=positivity,
        wave_peaks=wave_peaks,
        wave_resolution=wave_resolution,
        consistency=consistency,
        s3_all=s3_all,
        mh_all=mh_all,
        comparison_summary=comparison_summary,
        suppression_sensitivity=suppression_sens,
        s3_sensitivity=s3_sens,
        full_primary=full_primary,
        full_adult=full_adult,
    )

    if not args.no_figures:
        make_figures(s1, positivity, wave_peaks, s3_all, mh_all, output_dir)

    print("All manuscript-facing validation checks passed.")
    print(f"Outputs written to: {output_dir}")


if __name__ == "__main__":
    main()
```
