## Supplementary material for "The COVID-19 Trajectory in Israel: A Comprehensive Analysis of National Surveillance Data on Documented Infections, Test Positivity, and Vaccine-Effectiveness Estimates": README_Supplementary_Datasets.docx

**Supplementary National COVID-19 Surveillance Datasets S1-S5**
**README**

For the article: *The COVID-19 Trajectory in Israel: A Comprehensive Analysis of National Surveillance Data on Documented Infections, Test Positivity, and Vaccine-Effectiveness Estimates*

**Purpose**

This README describes the five supplementary datasets used in the longitudinal analysis of documented SARS-CoV-2 infection and vaccination in Israel. The files contain aggregate national surveillance data downloaded from the Israeli Ministry of Health (IMOH) dashboard/open-data system.

File preparation. For the supplementary release, Hebrew column headings in Datasets S1-S3 were translated into English. In Dataset S3, recurring age-group labels were also translated into English. Datasets S4-S5 retained the original English variable names supplied in the source files. No numerical values, dates, missing values, or privacy-suppressed values were altered.

**Study period**

Although Datasets S1-S3 extend through August 2, 2026, analyses were restricted to observations through May 27, 2023. The linked S4-S5 analysis used their common period, May 30, 2021 through May 27, 2023.

**Dataset overview**

| ID | Full data coverage | Role in study |
| --- | --- | --- |
| S1 | Feb 12, 2020-Aug 2, 2026 | Overall national trajectory of documented cases |
| S2 | Feb 20, 2020-Aug 2, 2026 | Overall national trajectory of test positivity |
| S3 | Jan 17, 2021-Aug 2, 2026 | Daily cases and rates per 100,000 by vaccination status |
| S4 | May 30, 2021-May 27, 2023 | Weekly cases by vaccination, age, and prior-infection status |
| S5 | Mar 15, 2020-May 27, 2023 | Weekly numbers tested by vaccination, age, and prior-infection status |

**Dataset S1**

File: Dataset_S1_Daily_Confirmed_Cases_Israel.xlsx

Worksheet: Daily Confirmed COVID-19 Cases

Content: Daily national counts of newly confirmed COVID-19 cases, the supplied 7-day moving average, and cumulative confirmed cases.

Study use: February 12, 2020-May 27, 2023; used to describe the overall epidemic trajectory.

| **Variable / field** | **Description** |
| --- | --- |
| Date | Calendar date. |
| Daily_confirmed_cases | Number of newly confirmed cases on the reported date. |
| 7_day_moving_average | Seven-day moving average of daily confirmed cases, as supplied in the source file. |
| Cumulative_confirmed_cases | Cumulative number of confirmed cases reported to date. |

**Dataset S2**

File: Dataset_S2_Daily_Test_Positivity_Israel.xlsx

Worksheet: Percent_tested_positive

Content: Daily percentage of tested individuals with a positive SARS-CoV-2 result, including PCR, institutional antigen, and overall positivity measures.

Study use: February 20, 2020-May 27, 2023; used with Dataset S1 to describe the national epidemic trajectory.

| **Variable / field** | **Description** |
| --- | --- |
| Date | Calendar date. |
| PCR_positivity_percent | Percentage positive for PCR testing. |
| Institutional_antigen_positivity_percent | Percentage positive for institutional antigen testing, where available. |
| Overall_positivity_percent | Overall percentage of tested individuals who were positive. |

**Dataset S3**

File: Dataset_S3_Daily_Cases_and_Rates_by_Vaccination_Status_Israel.xlsx

Worksheet: Daily Confirmed Cases and Rates

Content: Daily confirmed cases and corresponding rates per 100,000 population by vaccination status, reported for All ages, <60, and 60+ age groups.

Study use: January 17, 2021-May 27, 2023; used to reconstruct the vaccination-status-specific pattern visible in routine IMOH surveillance.

| **Variable / field** | **Description** |
| --- | --- |
| Date | Calendar date. |
| Age_group | Age category: All ages, <60, or 60+. |
| Cases_vaccinated | Confirmed cases among individuals classified as vaccinated. |
| Cases_expired_vaccination_status | Confirmed cases among individuals whose vaccination status was classified as expired. |
| Cases_unvaccinated | Confirmed cases among individuals classified as unvaccinated. |
| Cases_per_100k_vaccinated | Daily confirmed-case rate per 100,000 among vaccinated individuals. |
| Cases_per_100k_expired_vaccination_status | Daily confirmed-case rate per 100,000 among individuals with expired vaccination status. |
| Cases_per_100k_unvaccinated | Daily confirmed-case rate per 100,000 among unvaccinated individuals. |

**Dataset S4**

File: Dataset_S4_Weekly_Cases_by_Vaccination_Age_and_Recovery_Status_Israel.csv

Content: Weekly confirmed cases by age group, documented prior-infection (recovery) status, and vaccination status. The file also includes a total testing field for each week and age group.

Coverage: 104 calendar weeks, May 30, 2021-May 27, 2023; eight age groups (0-19, 20-29, 30-39, 40-49, 50-59, 60-69, 70-79, and 80+).

Variable names: Original English variable names were retained unchanged, including source spelling.

| **Variable / field** | **Description** |
| --- | --- |
| Week | Calendar week, represented as start date - end date. |
| Age_group | Age group. |
| Tests_num | Total testing field supplied in the source dataset for the corresponding week and age group. |
| Not_recovered_at_least_2_doses | Confirmed cases among persons without a documented previous infection who had received at least two vaccine doses. |
| Not_recovered_partially_vaccinated | Confirmed cases among persons without a documented previous infection who were partially vaccinated. |
| Not_recovered_not_vaccinated | Confirmed cases among persons without a documented previous infection who were unvaccinated. |
| Cases_among_recoverd_with_vacc | Confirmed cases among previously infected/recovered persons who were vaccinated. Variable name retained as supplied in the source file. |
| Cases_among_recovered_without_vacc | Confirmed cases among previously infected/recovered persons who were not vaccinated. |

**Dataset S5**

File: Dataset_S5_Weekly_Tests_by_Vaccination_Age_and_Recovery_Status_Israel.csv

Content: Weekly numbers of individuals tested, stratified by age group, documented prior-infection (recovery) status, and number of vaccine doses.

Coverage: 167 calendar weeks, March 15, 2020-May 27, 2023; ten age groups (0-9, 10-19, 20-29, 30-39, 40-49, 50-59, 60-69, 70-79, 80-89, and 90+).

Study use: For linkage with Dataset S4, analyses used the overlapping period from May 30, 2021 through May 27, 2023.

Variable names: Original English variable names were retained unchanged.

| **Variable / field** | **Description** |
| --- | --- |
| Test_week | Calendar week, represented as start date - end date. |
| Age_group | Age group. |
| Recovered_not_vaccinated | Previously infected/recovered individuals tested who were unvaccinated. |
| Recovered_1_dose | Previously infected/recovered individuals tested who had received one dose. |
| Recovered_2_doses | Previously infected/recovered individuals tested who had received two doses. |
| Recovered_3_and_more_doses | Previously infected/recovered individuals tested who had received three or more doses. |
| Non_recovered_not_vaccinated | Individuals without a documented previous infection who were tested and unvaccinated. |
| Non_recovered_1_dose | Individuals without a documented previous infection who were tested and had received one dose. |
| Non_recovered_2_doses | Individuals without a documented previous infection who were tested and had received two doses. |
| Non_recovered_3_and_more_doses | Individuals without a documented previous infection who were tested and had received three or more doses. |

**Linkage and harmonization used in the study**

Datasets S4 and S5 were linked by calendar week and harmonized age strata. The 0-9 and 10-19 groups in Dataset S5 were combined to correspond to the 0-19 group in Dataset S4. The 80-89 and 90+ groups in Dataset S5 were combined to correspond to the 80+ group in Dataset S4.

For the testing-informed vaccination-status comparison, analyses were restricted to individuals without a previous documented infection. The unvaccinated case category in Dataset S4 (Not_recovered_not_vaccinated) was matched to Non_recovered_not_vaccinated in Dataset S5. The vaccinated case category in Dataset S4 (Not_recovered_at_least_2_doses) was matched to the sum of Non_recovered_2_doses and Non_recovered_3_and_more_doses in Dataset S5. One-dose/partially vaccinated individuals were excluded from this comparison.

Privacy-suppressed counts. Cells reported as <5 were retained as <5 in the supplementary files. For analysis, these cells were treated as counts of 1-4 and assigned 2.5 in the primary calculation; sensitivity analyses used 1 and 4.

Further analytical transformations and statistical procedures are described in the manuscript Methods and accompanying analysis code.
